# Diagnostic delay of sarcoidosis: protocol for an integrated systematic review

**DOI:** 10.1101/2022.05.30.22275771

**Authors:** Tergel Namsrai, Jane Desborough, Dianne Gregory, Elaine Kelly, Matthew Cook, Christine Phillips, Anne Parkinson

**Author notes:** Correspondence Anne Parkinson, National Centre for Epidemiology and Population Health, Australian National University, 63, Eggleston Road, Acton ACT, 2601, Australia. **Funding** This review is part of the “Missed opportunities in clinical practice: Tools to enhance healthcare providers’ awareness and diagnosis of rare diseases in Australia” project funded by the Commonwealth represented by Department of Health Australia (Grant ID 4-G5ZN0T7). The funders had and will have no role in study design, data collection and analysis, decision to publish or preparation of the manuscript. **Competing interests** The authors have declared that no competing interests exist. **Data availability** No datasets were generated or analysed during the current study. All relevant data from this study will be made available upon study completion.

## Abstract

Sarcoidosis is a rare systemic inflammatory granulomatous disease with broad manifestation ranging from acute epileptic seizures to fatigue and pain syndromes that are subject to the organ involved. Delays in the diagnosis of sarcoidosis are attributed to the lack of a single diagnostic test or unified commonly used diagnostic criteria, and diagnosis based on exclusion of possible alternative diagnoses. We aim to systematically review the evidence about diagnostic delay in sarcoidosis to elucidate the causes and consequences of diagnostic delay, including people with sarcoidosis’ experiences. This will inform the development of interventions, tools, and health policies aiming to improve diagnostic efficiency and patients’ experiences of sarcoidosis.

**Methods and analysis:** A systematic search of the literature will be conducted using PubMed/Medline, Scopus, and ProQuest databases, and sources of grey literature, up to 25^th^ of May 2022, with no limitations on publication date. We will include all study types (qualitative, quantitative, and mixed methods) except review articles, examining diagnostic delay, incorrect diagnosis, missed diagnosis or slow diagnosis of all types of sarcoidosis across all age groups. We will also examine evidence of patients’ experiences associated with diagnostic delay. Only studies in English, German and Indonesian will be included. The outcomes we examine will be diagnostic delay time, patients’ experiences, and causes and consequences associated with diagnostic delay in sarcoidosis. Two people will independently screen the titles and abstracts of search results, and then the remaining full-text documents against the inclusion criteria. Disagreements will be resolved with a third reviewer until consensus is reached. Selected studies will be appraised using the Mixed Methods Appraisal Tool (MMAT). A meta-analysis and subgroup analysis of quantitative data will be conducted. Meta-aggregation methods will be used to analyse qualitative data. If there is insufficient data for these analyses, a narrative synthesis will be conducted.

**Ethics and dissemination:** Ethical approval will not be required as no human recruitment or participation will be involved. Findings of the study will be disseminated through publications in peer-reviewed journals, conferences, and symposia.

**Trial registration:** PROSPERO Registration number: CRD42022307236 URL of the PROSPERO registration: https://www.crd.york.ac.uk/PROSPEROFILES/307236_PROTOCOL_20220127.pdf

## Introduction

Sarcoidosis is a rare systemic inflammatory granulomatous disease of unknown cause with broad manifestation ranging from acute epileptic seizures to subtle symptoms such as fatigue and pain; syndromes that are subject to the organ involved (1-3). Sarcoidosis can manifest in any organ including the lungs, skin, liver, joints, nervous system, and eyes (4, 5), but it most commonly affects the lungs, referred to as pulmonary sarcoidosis (PS) (6). There is no single diagnostic test for sarcoidosis or a unified, commonly used diagnostic criteria. Diagnosis of sarcoidosis relies on clinical manifestations along with radiological or histological evidence and exclusion of possible alternative diagnoses (7).

Due to its rarity, broad range of clinical features, lack of conclusive diagnostic testing and the requirement for diagnosis to be based on exclusion of other possible diagnoses, timely diagnosis of sarcoidosis can be challenging and result in substantial diagnostic delays and inappropriate treatment. This can potentially result in people with sarcoidosis living with pain and other unnecessary symptoms until the correct diagnosis is determined. Research examining diagnostic delay, including factors associated with diagnostic delay and people’s experience of diagnostic delay in sarcoidosis, is rare. Understanding factors associated with diagnostic delay and people’s experiences of this could inform the development of interventions aimed at enhancing diagnostic efficiency and improving people’s experiences of living with sarcoidosis.

## Objective

The aim of this integrated systematic review is to review the evidence regarding diagnostic delay in sarcoidosis. To this end, our aim is to answer two key research questions:

RQ1. What are the causes and consequences of diagnostic delay of sarcoidosis?

RQ2. What evidence is there about patients’ experience of sarcoidosis’ diagnostic delay?

## Methods and Analysis

### Protocol development

This study protocol has been developed in accordance with the Preferred Reporting Items for Systematic Review and Meta-Analysis Protocols (PRISMA-P) and the Cochrane Handbook for Systematic Reviews (8, 9).

### Search strategy

The search strategy was developed to ensure reproducibility and increase transparency following the PRISMA-P checklist (8). Research questions and search terms were developed using the PICOS tool (Population/Intervention/Comparison/Outcomes/Study Design) to ensure reliability and homogeneity of search results (10). The study is registered with PROSPERO (CRD42022289830). A systematic search of peer reviewed literature will be conducted using PubMed/Medline, Scopus, and ProQuest databases, and searches of the grey literature will include Open Access Theses and Dissertation (https://oatd.org/), ProQuest Thesis and Dissertations and the National Library of Australia. Reference lists of selected studies and review articles will also be searched.

Search terms were developed in collaboration with research team members (**, **, **), and combined using Boolean operators “AND” and “OR”. A preliminary exploratory search on PUBMED/MEDLINE was conducted on 15^th^ October 2021 (Table 1) to inform the final search strategy and determine outcomes. This search strategy was updated and peer reviewed (**, **) using the PRESS checklist (11). The final search terms will include sarcoidosis AND (“delay in diagnosis” OR “diagnostic delay” OR “misdiagnosis” OR “time to diagnosis” OR “incorrect diagnosis” OR “missed diagnosis” OR “delayed diagnosis”) without restrictions on study type, date, and language. The literature search results will be imported to Covidence, an internet-based software that facilitates collaboration between reviewers and ensures independent review of the literature (12).

**Table 1.** Search string conducted on PUBMED/MEDLINE

| Table 1. Search string conducted on PUBMED/MEDLINE |  |  |
| --- | --- | --- |
| Search number | Query | Search Details |
| 1 | sarcoidosis [Title/Abstract] | sarcoidosis [Title/Abstract] |
| 2 | delay in diagnosis<br>[Title/Abstract] | delay in diagnosis [Title/Abstract] |
| 3 | delayed diagnosis<br>[Title/Abstract] | delayed diagnosis [Title/Abstract] |
| 4 | diagnostic delay<br>[Title/Abstract] | diagnostic delay [Title/Abstract] |
| 5 | time to diagnosis<br>[Title/Abstract] | time to diagnosis [Title/Abstract] |
| 6 | misdiagnosis [Title/Abstract] | misdiagnosis [Title/Abstract] |
| 7 | missed diagnosis<br>[Title/Abstract] | missed diagnosis [Title/Abstract] |
| 8 | incorrect diagnosis<br>[Title/Abstract] | incorrect diagnosis [Title/Abstract] |
| 9 | #2 OR #3 OR #4 OR #5 OR<br>#6 OR #7 OR #8 | delay in diagnosis [Title/Abstract] OR<br>"delayed diagnosis"[Title/Abstract] OR<br>"diagnostic delay"[Title/Abstract] OR "time<br>to diagnosis"[Title/Abstract] OR<br>"misdiagnosis"[Title/Abstract] OR "missed<br>diagnosis"[Title/Abstract] OR "incorrect<br>diagnosis"[Title/Abstract] |
| 10 | #1 AND #9 | sarcoidosis [Title/Abstract] AND ("delay in<br>diagnosis"[Title/Abstract] OR "delayed<br>diagnosis"[Title/Abstract] OR "diagnostic<br>delay"[Title/Abstract] OR "time to<br>diagnosis"[Title/Abstract] OR<br>"misdiagnosis"[Title/Abstract] OR "missed<br>diagnosis"[Title/Abstract] OR "incorrect<br>diagnosis"[Title/Abstract]) |

### Study selection

Studies will be selected according to the pre-developed PICOS eligibility criteria outlined in Table 2. All study types (qualitative, quantitative, and mixed methods) except review articles, examining diagnostic delay, incorrect diagnosis, missed diagnosis or slow diagnosis of all types of sarcoidosis will be included. We will also examine evidence of patients’ experiences associated with diagnostic delay. Given the nature of the study there will be no comparison group. No setting or publication date limitations will be applied. Only studies in English, German and Indonesian will be included.

**Table 2.** Inclusion and exclusion criteria

| Table 2. Inclusion and exclusion criteria |  |  |
| --- | --- | --- |
| PICOS | Inclusion criteria | Exclusion criteria |
| Population | Studies examining people with sarcoidosis of all ages | - |
| Intervention/Exposure | Studies examining delayed, incorrect diagnosis, missed diagnosis or slow diagnosis of sarcoidosis | - |
| Comparison | Not applicable | - |
| Outcome | Primary outcome: diagnostic delay.<br>Secondary outcomes:<br>i) causes and consequences of diagnostic delay | - |
|  | ii) people with sarcoidosis'<br>experiences of diagnostic delay |  |
| Study design | All study designs | Review articles |
| Language | English, German, Indonesian |  |
| Setting | No restriction | - |
| Timing | No restriction | - |

As a first step, the title and abstract of the literature search results will be independently screened by two review authors (** and **), against the pre-developed inclusion criteria. Any conflicts will be discussed among the review team and resolved by a third reviewer (**).

Full texts will be sourced for all studies that meet the inclusion criteria or where there is any uncertainty. Two review authors (** and **) will then screen the full text documents according to the inclusion criteria, with any conflicts resolved by a third reviewer (**). Exclusion rationales will be recorded.

### Data extraction

Following completion of the study selection process, a data extraction tool will be designed, peer reviewed and piloted. In the piloting process, two reviewers (** and **) will independently extract data from the same five studies and compare their results to establish consensus and validity of the data extraction tool.

Data items to be extracted include:

1. Identification of the study (journal, authors, year, citation, research center/university/hospital/organisation, conflict of interest, funding/sponsorship),
2. Methods (study aim, study design, participant demographics, recruitment process, inclusion, exclusion criteria, statistical analysis),
3. Main findings (exposure details, diagnostic delays, causes and consequences of delay, patients’ experience, and other relevant outcomes).

In cases of missing information about diagnostic delay, when available, the date of symptom onset and date of diagnosis will be used to calculate diagnostic delay. Any disagreements will be resolved through discussion, conflicts will be resolved by a third reviewer (**). Study authors will be contacted to resolve uncertainties about extracted data.

The primary outcome of the review is diagnostic delay time (time from symptom onset to correct diagnosis) in people living with sarcoidosis. Secondary outcomes include patients’ experiences, causes and consequences of diagnostic delay in sarcoidosis.

### Quality appraisal

The Mixed Methods Appraisal Tool (MMAT), will be used to appraise the quality of included studies (quantitative, qualitative and mixed methods) (13). If the selected studies are only quantitative, an appropriate adapted version of Newcastle-Ottawa scale (14) will be used depending on the study types included. The chosen quality appraisal tool will be piloted by two independent review authors (** and **), on a randomly selected five samples with any conflicts resolved by a third reviewer following discussion (**). An independent reviewer (**) will continue quality appraisal on the remaining studies.

### Data synthesis and meta-analysis

A systematic narrative synthesis will be undertaken to explore the findings of included studies in relation to time from symptom onset to diagnosis, and people’s experiences related to delayed diagnosis in line with guidance from the Centre for Reviews and Dissemination (15).

A meta-analysis will also be conducted if extracted quantitative data are homogenous, using a random-effects model in conjunction with subgroup analyses, including pooled diagnostic delay data in each type of sarcoidosis. Extracted qualitative data will be meta-synthesized using meta-aggregation. Similarly, processed data (findings) from qualitative studies will be extracted and aggregated into a single set of categories, which will then be further aggregated and synthesised into a set of statements that may be useful to inform clinical practice.

### Quality of evidence

The quality/certainty of evidence for all quantitative outcomes included in a meta-analysis will be judged using the Grading of Recommendations Assessment, Development and Evaluation (GRADE) working group methodology (16). The domains of risk of bias, consistency of effect, imprecision, indirectness, and publication bias will be used to assess the certainty of the body of evidence, which will be reported in four levels: high, moderate, low, and very low.

### Amendments

If the protocol is amended prior to commencing the study, these amendments (date, explanation, and rationale) will be described in the final protocol. The record will be in tabular format as recommended by the Cochrane Collaboration (8).

## Data Availability

No datasets were generated or analysed during the current study. All relevant data from this study will be made available upon study completion.

https://www.crd.york.ac.uk/PROSPEROFILES/307236_PROTOCOL_20220127.pdf

## Author contributions

Conceptualization: Anne Parkinson, Jane Desborough, Dianne Gregory, Elaine Kelly, Matthew Cook, Christine Phillips Data curation: Tergel Namsrai, Anne Parkinson, Jane Desborough Investigation: Tergel Namsrai, Anne Parkinson, Jane Desborough Methodology: Tergel Namsrai, Anne Parkinson, Jane Desborough, Matthew Cook, Christine Phillips Project coordinator: Anne Parkinson Writing – original draft: Tergel Namsrai Writing – review & editing: Anne Parkinson, Jane Desborough, Dianne Gregory, Elaine Kelly, Matthew Cook, Christine Phillips

## Supplementary files

Original registered protocol is attached as Supplementary file 1.

